# Non-attendance in Telephone versus In-person Secondary Care Consultations: Retrospective cohort Study of Patients with Type 2 Diabetes in Northwest London

**DOI:** 10.64898/2026.05.07.26352666

**Authors:** Reham Aldakhil, Geva Greenfield, Gabriele Kerr, Benedict Hayhoe, Holger Kunz, Jonathan Valabhji, Azeem Majeed, Ana Luisa Neves

## Abstract

**Background:** Although virtual consultations are increasingly used in healthcare, mode affects attendance patterns remains limited, particularly across demographic groups. Within NHS secondary care, telephone consultations have been the most widely adopted form of telephone care; however, few studies have examined non-attendance (commonly termed ‘Did Not Attend’ [DNA]) patterns specifically for telephone consultations and fewer still have explored how patient characteristics influence attendance differently across consultation modes. Understanding these patterns is essential for equitable service planning.

**Objective:** To compare non-attendance rates between telephone and in-person secondary care consultations among adults with type 2 diabetes (T2D), and to identify patient characteristics associated with non-attendance under each mode.

**Methods:** Data from 853,693 secondary care consultations (January 2020-August 2024) for 45,618 patients with T2D in Northwest London were analysed. Telephone consultations in this dataset consisted exclusively of telephone consultations; we therefore refer to them as ‘telephone consultations’ throughout. Patient-level consultations were aggregated into patient-mode strata for regression modelling. Zero-inflated Negative Binomial regression assessed factors associated with missed consultation rates by mode (in-person or telephone). Propensity-score balance diagnostics (inverse probability of treatment weighting) were conducted to assess measured confounding by mode assignment. Specialty-stratified non-attendance rates were examined across 34 major specialties.

**Results:** In-person consultations had higher unadjusted non-attendance rates than telephone consultations (9.1% vs 7.2%, p<0.001). This pattern was consistent for both first consultations (9.3% vs 6.2%, p<0.001) and follow-up consultations (9.0% vs 7.50%, p<0.001). For in-person consultations, higher non-attendance was associated with younger age (18-39: 12.2%, 40-59: 11.1% vs 60-79: 9.9%, p<0.001), Black or Black British ethnicity (18.9% vs White: 7.6%, p<0.001), and greater deprivation (most deprived IMD1: 10.3% vs least deprived IMD5: 7.0%, p<0.001). For telephone consultations, higher non-attendance was associated with male gender (7.3% vs female: 7.0%, p<0.01), younger age (18-39: 11.3%, 40-59: 9.5% vs 60-79: 6.1%, 80+: 5.6%, p<0.001), and greater socioeconomic deprivation (most deprived: 8.3% vs least deprived: 4.7%, p<0.001). Interaction analyses revealed that demographic disparities were amplified for telephone relative to in-person consultations. Specialty-stratified analysis showed that in-person non-attendance exceeded telephone non-attendance in the majority of high-volume specialties.

**Conclusions:** In-person consultations had higher non-attendance rates than telephone consultations. Various demographic factors influenced non-attendance rates, with younger age and greater socioeconomic deprivation consistently associated with non-attendance for both in-person and telephone consultations. These findings suggest that a personalised, equity-informed approach to consultation mode selection is needed. Findings apply to telephone consultations and may not generalise to video-based modalities.

## Background

The complexity of long-term conditions care delivery, coupled with the contemporary challenges experienced by healthcare systems (i.e., workforce shortages, increasing complex care, and increasing need for care integration), has necessitated new approaches to care delivery[1]. One example has been the increasing use of telephone consultations (VC), either as a complement or as an alternative to traditional in-person consultations [2]. Telephone consultations offer potential advantages over in-person care provision in improving healthcare accessibility and reducing treatment burdens for patients [3]. Telephone care can decrease travel time, reduce work absences, and potentially improve consultation attendance [4–6]. Within NHS secondary care, telephone consultations have been the most widely adopted form of virtual care to date, and their attendance patterns may differ from video-based alternatives. However, digital literacy, availability of technology, language barriers, and socioeconomic factors can negatively impact patients’ willingness and ability to engage with telephone consultations. This might be reflected in reduced engagement with these technologies and lower attendance rates [7, 8]. Uptake of such technologies can vary across different patient groups, potentially exacerbating existing inequities in healthcare services delivery [4, 5, 9]. However, limited evidence exists on how patient characteristics influence attendance at telephone versus in-person consultations [5, 10, 11]. Understanding attendance of telephone consultations and exploring variations based on patient characteristics is crucial for assessing uptake and ensuring these new care models are equitable for diverse patient populations.

Type 2 diabetes (T2D) is a complex condition, often co-existing with other long-term conditions. While primary care manages most routine diabetes care, people with T2D often require specialist input from various secondary care services, including endocrinology, cardiology, ophthalmology, nephrology, and podiatry due to the multi-system complications associated with the disease [12]. The cost of T2D care to the NHS in the UK is estimated at £10 billion per year, of which approximately £3 billion is attributed to specialist outpatient care [13]. T2D therefore represents an important exemplar of a long-term condition for which telephone consultations in a range of secondary care specialties may have significant impact. Exploring engagement with telephone consultations in this context is therefore of great importance for planning of care pathways.

We compared non-attendance rates in telephone versus in-person secondary care outpatient consultations for individuals with T2D; and examined which patient characteristics associated with non-attendance in in-person and telephone.

## Methods

### Data source and variables

Data were sourced from the Whole Systems Integrated Care (WSIC) database, which links coded data for over 2.7 million patients registered with GP practices in Northwest London. The dataset includes primary and secondary care and is linked to Emergency Department (ED) records, outpatient department attendances, and inpatient admissions [14]. The retrospective cohort included all patients diagnosed with type 2 diabetes before 2020 who had at least one scheduled telephone or in-person consultation between January 2020 and August 2024.

Non-attendance (commonly termed ‘Did Not Attend’ [DNA] in NHS administrative data) was defined as consultations not attended by patients without being cancelled in advance. For telephone consultations, non-attendance means the patient did not answer the call or engage with the remote session. The WSIC database records the consultation mode actually delivered but does not capture the clinical, service-level, or patient-level reasons for offering a telephone versus an in-person consultation in any individual case. Mode assignment during the study period reflected a combination of clinician discretion, specialty-specific service redesign, pandemic-era service constraints, and patient preference; none of these determinants are systematically available in the routine dataset.

A low proportion of missing data was observed (<5% for all variables; gender 0%, age 1.8%, ethnicity 4.2%, IMD quantile 0.8%, and consultation mode 2.4%). To assess whether missing data might bias our results, patterns of missingness across variables were examined and characteristics between cases with complete data and those with missing values were compared. No significant differences were found in demographic characteristics between complete and incomplete records. A detailed overview of the missing data analysis is provided in **Error! Reference source not found**.. Given the low proportion and balanced pattern of missingness, complete case analysis was employed. Records with ‘Unknown/Not stated’ ethnicity were retained as an explicit missing-indicator category rather than excluded. A detailed overview of the missing data analysis is provided in Supplementary File 1.

The consultation mode (in-person or telephone), type (first or follow-up), and attendance status are defined by NHS Digital, established and standardised as part of the processing cycle and data quality checks applied to commissioning datasets [15, 16]. Virtual consultations in this dataset consisted exclusively of telephone consultations. A detailed description of the outcome and predictor variables is provided in Textbox 1.

**Textbox 1.**
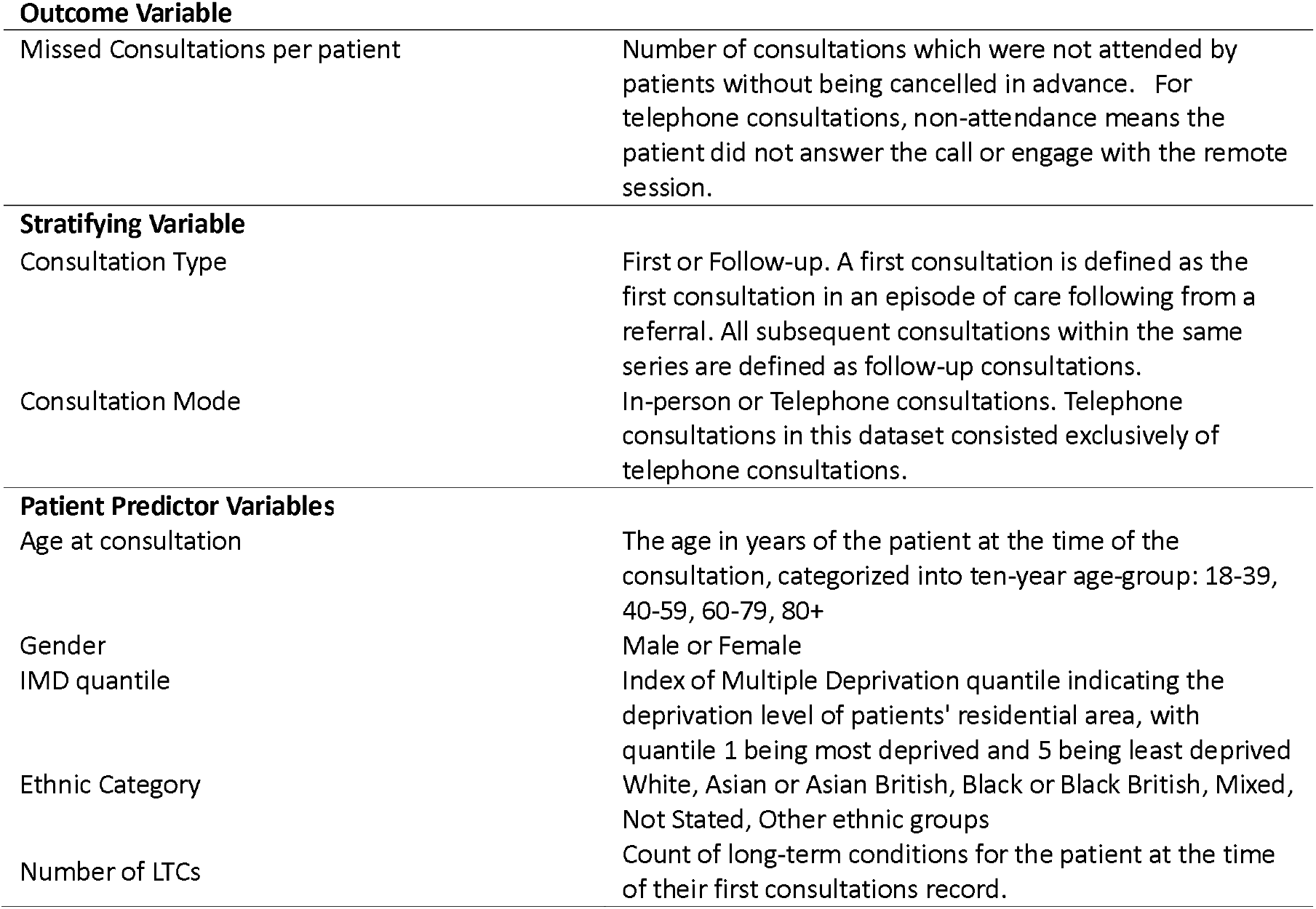
Variables Used in the analysis of Non-attendance in T2D Secondary Care Consultations.

**Outcome Variable**
|  |  |
| --- | --- |
| Missed Consultations per patient | Number of consultations which were not attended by patients without being cancelled in advance. For telephone consultations, non-attendance means the patient did not answer the call or engage with the remote session. |
| **Stratifying Variable** |  |
| Consultation Type | First or Follow-up. A first consultation is defined as the first consultation in an episode of care following from a referral. All subsequent consultations within the same series are defined as follow-up consultations. |
| Consultation Mode | In-person or Telephone consultations. Telephone consultations in this dataset consisted exclusively of telephone consultations. |
| **Patient Predictor Variables** |  |
| Age at consultation | The age in years of the patient at the time of the consultation, categorized into ten-year age-group: 18-39, 40-59, 60-79, 80+ |
| Gender | Male or Female |
| IMD quantile | Index of Multiple Deprivation quantile indicating the deprivation level of patients' residential area, with quantile 1 being most deprived and 5 being least deprived |
| Ethnic Category | White, Asian or Asian British, Black or Black British, Mixed, Not Stated, Other ethnic groups |
| Number of LTCs | Count of long-term conditions for the patient at the time of their first consultations record. |

### Statistical Analysis

To compare unadjusted non-attendance rates between telephone and in-person consultations, chi-square tests were performed. At the consultation level, a total of 850,493 consultations were identified. These were aggregated into patient-mode strata for regression modelling.

To examine the association between patient characteristics and non-attendance, a Zero-Inflated Negative Binomial (ZINB) regression models were fitted. This approach was selected to account for both overdispersion in count data and excess zeros (representing patients who never miss consultations). Each ZINB model includes two components: (1) a count component modelling the expected rate of missed consultations per scheduled consultation, with an offset of log(total scheduled consultations per stratum), so that coefficients are interpretable as adjusted rate ratios (aRRs); and (b) a zero-inflation component modelling the probability of being ‘perfect attendaer’ (structural zero), with coefficients reported as adjusted odds ratios (aORs). The count component captures variation in missed-consultation rates among the population susceptible to missing, while the zero-inflation component captures the subpopulation of ‘always-attenders’ who never miss consultations regardless of other characteristics.

Predictors included age group (18–39 [reference], 40–59, 60–79, 80+), gender (female [reference], male), ethnicity (Asian or Asian British [reference], white, Black or Black British, Mixed, Other ethnic groups, Unknown), IMD quantile (quantile 1 most deprived [reference] through quantile 5 least deprived), and number of long-term conditions (1 [reference], 2, 3+), and appointment type (first [reference], follow-up). Model diagnostics confirmed the appropriateness of the ZINB specifications: was appropriate through residual plots, Vuong tests, and assessment of overdispersion. Detailed information on statistical assumptions, diagnostics, and model evaluation are provided in Supplementary File 3.

To formally test whether the association between each patient characteristic and non-attendance differed by mode, a joint ZINB model was fitted including mode × covariate interactions (mode × age, mode × gender, mode × ethnicity, mode × IMD, mode × number of LTCs; theta=0.45, converged in 84 iterations). Additionally, within-mode interaction models were fitted with gender × age group and ethnicity × IMD quantile interactions to examine patterns within each consultation mode.

To address confounding by non-random mode assignment, a propensity score for mode (telephone versus in-person) was estimated using logistic regression on age, gender, ethnicity, IMD quantile, LTC count, and specialty. Stabilised inverse probability of treatment weights (IPTW) were computed and covariate balance was assessed via standardised mean differences (SMDs) before and after weighting. In addition to relative measures, unadjusted absolute risk differences (ARDs) between modes are reported. Specialty-stratified non-attendance rates were examined across all major specialties to assess whether the overall pattern was consistent across service lines.

Analyses were conducted using R version 4.3.0 (R Foundation for Statistical Computing, Vienna, Austria) (Figure 1) [17].

**Figure 1.**
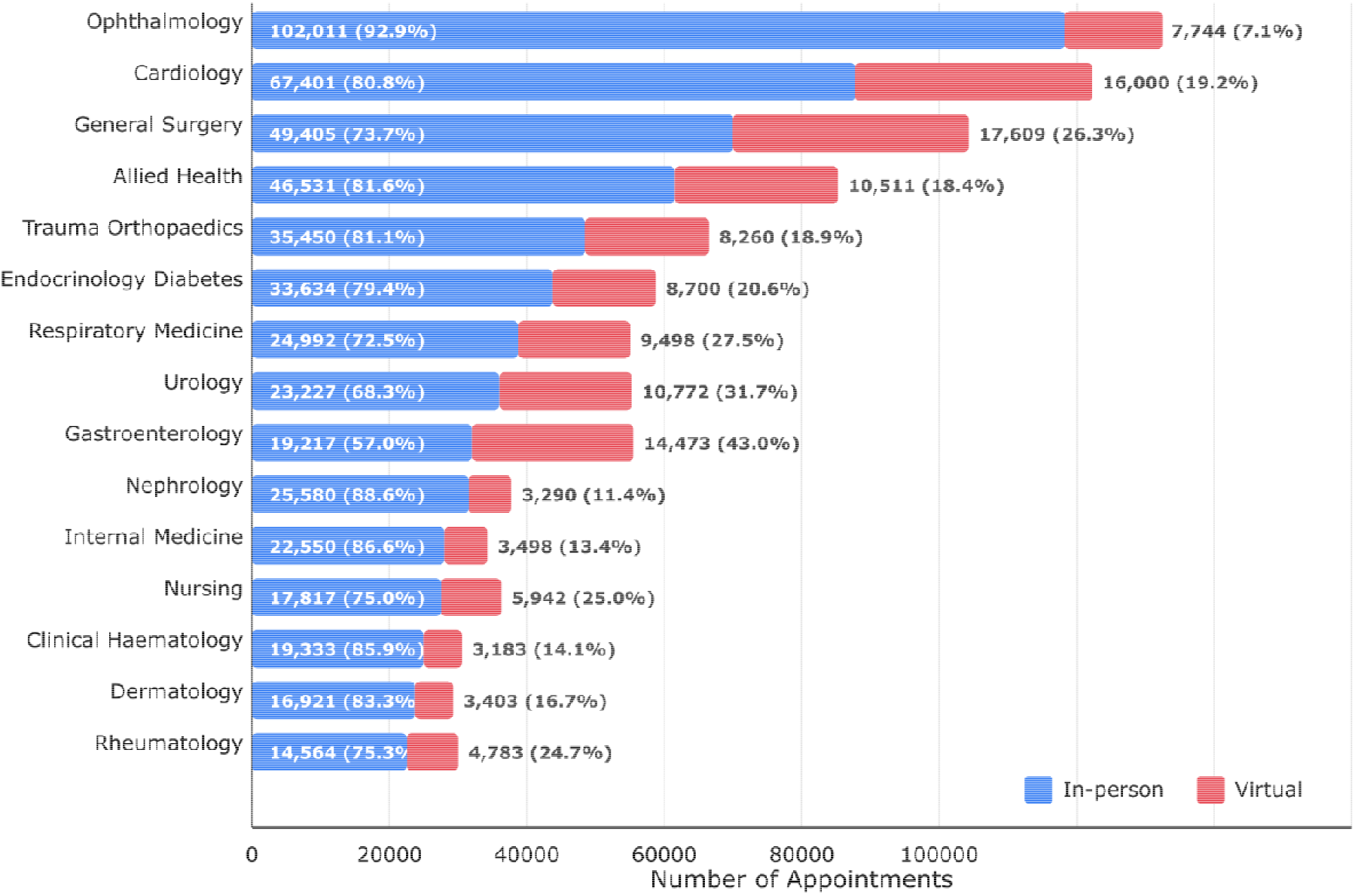
Secondary care specialty distribution by consultation mode. Bar chart showing the number of in-person (blue) and telephone (red) consultations across 15 medical specialties for patients with type 2 diabetes in Northwest London (January 2020-August 2024). Numbers and percentages within bars represent consultation counts and proportion of total consultations for each specialty. Ophthalmology had the highest proportion of in-person consultations (92.9%), whilst Gastroenterology showed the highest adoption of telephone consultations (43.0%). Total consultations analysed: 853,693 (80.8% in-person, 19.2% telephone).

### Data Availability

The data that support the findings of this study are not publicly available. Requests to access to the data sets used in this paper via a secure environment can be made via the Whole Systems Integrated Care (WSIC) Data Access Committee, subject to relevant approvals and data governance. Access requires institutional agreements and cannot be shared publicly.

### Ethics Approval

This study used fully pseudonymised, routinely collected data from the WSIC database, accessed under the WSIC information governance framework. The analysis was approved through the WSIC Data Access Committee review process and conducted within the WSIC secure analytics environment. Data access and governance procedures conformed to NHS England and Imperial College London institutional policies for use of anonymised electronic health records.

## Results

### Patient Characteristics

The study included 45,618 adults with type 2 diabetes. Most participants were male (52%, n= 23,754), with a mean age of 59.5 years. The ethnic distribution was diverse; the predominant ethnic group was Asian or Asian British subjects (47.7%, n=21,763), followed by White (26.3%, n=12,006), Black or Black British (13.1%, n=5,960). The ‘Other ethnic groups’ category (7.1%, n=3,243) comprised Chinese, Arab, and any other ethnic group as classified by the NHS Digital 2001 Census categories. More than half of patients had three or more long-term conditions (58.1%, n=26,481), and the majority lived in areas with moderate deprivation (IMD 2 and 3, together accounting for 63.9%). An overview of patient characteristics is provided in Table 1.

**Table 1.** Patient characteristics according to mode of consultation.

| Characteristic | In-person<br>Consultations n(%) | Telephone<br>Consultations n(%) | Total Consultations n(%) | p-value* |
| --- | --- | --- | --- | --- |
| Gender |  |  |  |  |
| Male | 362,040 (52.5) | 84,000 (51.3) | 446,040 (52.2) | p <0.001 |
| Female | 327,875 (47.5) | 79,778 (48.7) | 407,653 (47.8) |  |
| Age Group |  |  |  |  |
| 18 to 39 | 22,332 (3.2) | 4,544 (2.8) | 26,876 (3.1) | p<0.001 |
| 40 to 59 | 203,747 (29.5) | 48,159 (29.4) | 251,906 (29.5) |  |
| 60 to 79 | 381,155 (55.3) | 91,238 (55.7) | 472,393 (55.3) |  |
| 80+ | 82,023 (11.9) | 19,628 (12.0) | 101,651 (11.9) |  |
| Ethnicity |  |  |  |  |
| Asian or Asian British | 286,892 (41.6) | 71,161 (43.4) | 358,053 (41.9) | p<0.001 |
| White | 219,023 (31.8) | 50,579 (30.9) | 269,602 (31.6) |  |
| Black or Black British | 91,661 (13.3) | 21,091 (12.9) | 112,752 (13.2) |  |
| Other Ethnic Groups | 49,197 (7.1) | 11,583 (7.1) | 60,780 (7.1) |  |
| Mixed | 21,491 (3.1) | 4,999 (3.0) | 26,490 (3.1) |  |
| Unknown | 21,651 (3.1) | 4,365 (2.7) | 26,016 (3.0) |  |
| Long term conditions |  |  |  |  |
| 1 | 81,302 (11.8) | 19,191 (11.7) | 100,493 (11.8) | p=0.001 |
| 2 | 127,472 (18.5) | 29,746 (18.2) | 157,218 (18.4) |  |
| 3+ | 481,141 (69.7) | 114,841 (70.1) | 595,982 (69.8) |  |

| IMD quantile |  |  |  |  |
| --- | --- | --- | --- | --- |
| 1 (most deprived) | 113,388 (16.4) | 25,224 (15.4) | 138,612 (16.2) | p<0.001 |
| 2 | 239,798 (34.8) | 54,844 (33.5) | 294,642 (34.5) |  |
| 3 | 198,170 (28.7) | 46,911 (28.6) | 245,081 (28.7) |  |
| 4 | 99,819 (14.5) | 25,594 (15.6) | 125,413 (14.7) |  |
| 5 (least deprived) | 38,740 (5.6) | 11,205 (6.8) | 49,945 (5.9) |  |

### Consultation description

A total of 853,693 consultations were analysed. Of these, 689,915 (80.8%) were in-person and 163,778 (19.2%) were telephone. Statistical analysis showed significant differences in the demographic distribution between consultation modes (*P*<.001 for gender, age group, ethnicity and IMD quintile; *P*=0.001 for long term conditions). There was a slightly higher representation of Asian or Asian British patients in telephone consultations (43.4% vs 41.6% for in-person) and patients from the least deprived areas (IMD quantile 5: 6.8% in telephone vs 5.6% in in-person).

Consultations were distributed across various secondary care specialties. Ophthalmology represented the highest volume of consultations accounting for 12.9% of all consultations (n=109,755, 92.9% in person, 7.1% telephone), followed by Cardiology (9.8% of all consultations, n=83,401, 80.8% in person, 19.2% telephone), and General Surgery (7.8% of all consultations, 73.7% in person, 26.3% telephone). While most specialties maintained predominantly in-person visits, certain specialties demonstrated higher adoption of telephone consultations, with Gastroenterology showing the highest proportion of telephone consultations at 43.0% (n=14,473) of its total consultations (Figure 1).

### Non-attendance rates in telephone and in-person consultations

Overall, in-person consultations showed a higher non-attendance than telephone consultations (9.1% vs 7.2%, absolute risk difference 2.0 percentage points). This pattern was consistent by consultation type, with in-person consultations showing higher non-attendance than telephone for both first consultations (9.3% vs 6.2%, *P*<.001) and follow-up consultations (9.0% vs 7.5%, *P*<.001).

### Determinants of non-attendance in in-person and telephone consultations

All effect sizes reported below are adjusted rate ratios (aRRs) from the count component of stratified ZINB models, unless otherwise specified. An overview is provided in Figure 2. Detailed information of the regression model is provided in **Error! Reference source not found**..

**Figure 2.**
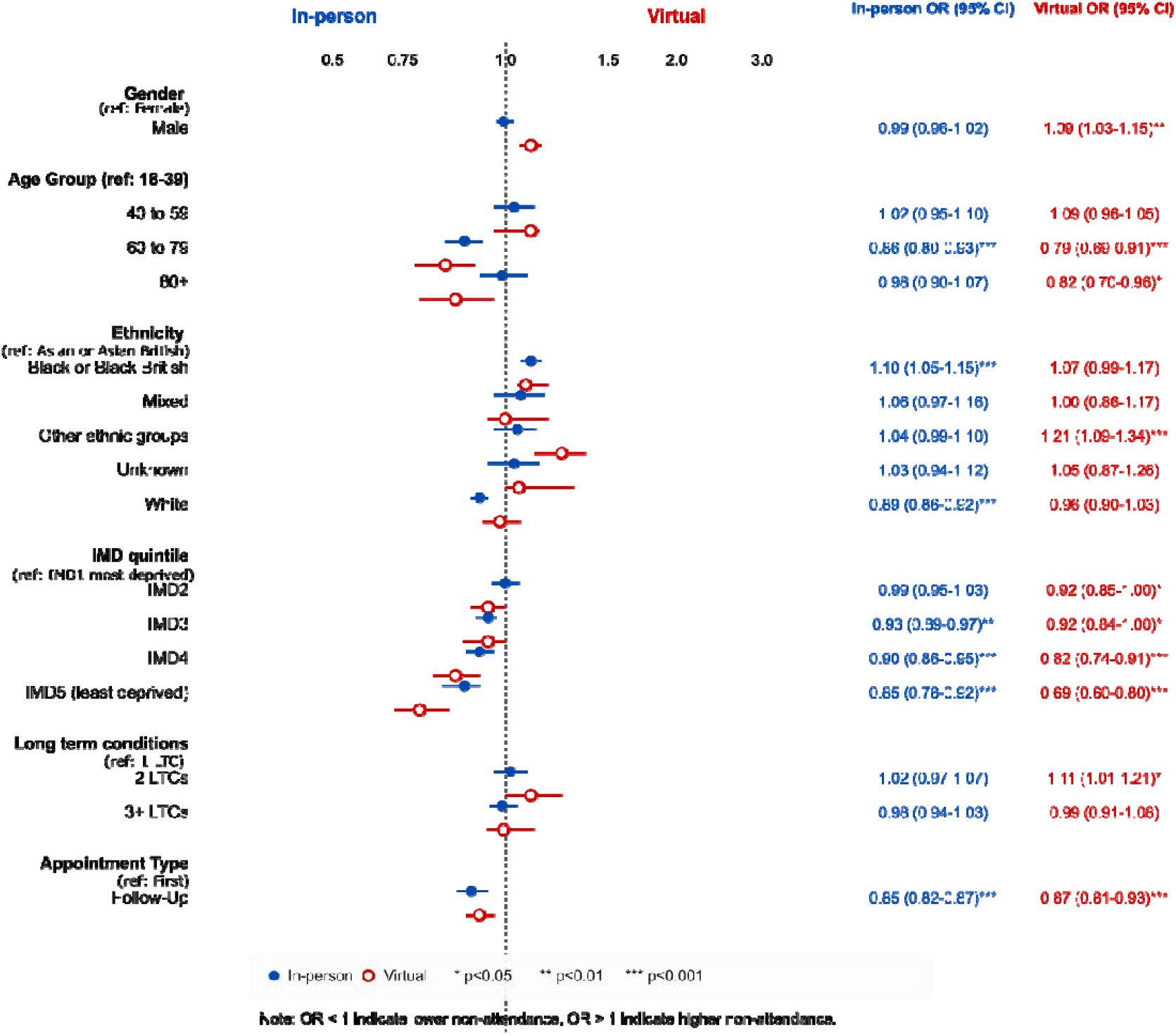
Factors associated with non-attendance in in-person and telephone consultations. Odds ratios (ORs) with 95% confidence intervals for non-attendance at in-person (blue) versus telephone (red) healthcare consultations by demographic and clinical characteristics. Reference categories are indicated in parentheses for each variable group. ORs < 1 indicate lower probability of non-attendance compared to the reference group, while ORs > 1 indicate higher probability.

For in-person consultations, younger patients (18-39 years) had higher non-attendance rates (12.2%) compared to older patients aged 60-79 (9.9%). Black or Black British patients had significantly higher non-attendance rates (18.9%) compared to the Asian or Asian British reference group (8.7%; *P*<.001). A clear socioeconomic gradient was observed, with patients from more deprived areas (IMD quintile 1) showing the highest non-attendance rate. Patients from the most deprived areas (IMD quantile 1) had the highest non-attendance (10.3%), compared to the least deprived areas (IMD5: 7.0%, *P*<.001). Detailed estimates are presented in Figure 2 and Supplementary File 2.

For telephone consultations, male patients showed higher non-attendance (7.3%, vs female: 7.0%; P<.01). Younger patients also had higher non-attendance with those aged 18-39 showing the highest rates (11.3%), while older patients aged 60-79 (6.1%) and 80+ (5.6%,) had significantly lower non-attendance rates (*P*<.001). Patients from Other ethnic groups had higher non-attendance (8.6%, P<.001), and a socioeconomic gradient was also observed (IMD1: 8.3% vs IMD5: 4.7%; P<.001). Full results are in Supplementary File 2.

### Interaction Effects

To examine differential effects of patient characteristics on consultation non-attendance by mode, the interaction terms between consultation mode and patient factors were analysed **Error! Reference source not found**..

Two principal patterns emerged from the interaction models (Figure 3). First, young men (18–39 years) had markedly higher non-attendance than young women under both modes, with the gender gap narrowing in older age groups (male × age 60–79 interaction: aOR=0.70, P<.001). Second, the ethnicity × deprivation interaction showed that ‘Other ethnic groups’ in the least deprived areas (IMD5) had notably high non-attendance (aOR=1.58, P<.001). The joint ZINB model with mode × covariate interactions confirmed that demographic disparities were amplified for telephone relative to in-person consultations (age 80+ × Remote: aOR=2.65, 95% CI 2.45–2.88; Male × Remote: aOR=1.64, 95% CI 1.59–1.69). Full interaction estimates are in Figure 3 and Supplementary File 2.

**Figure 3.**
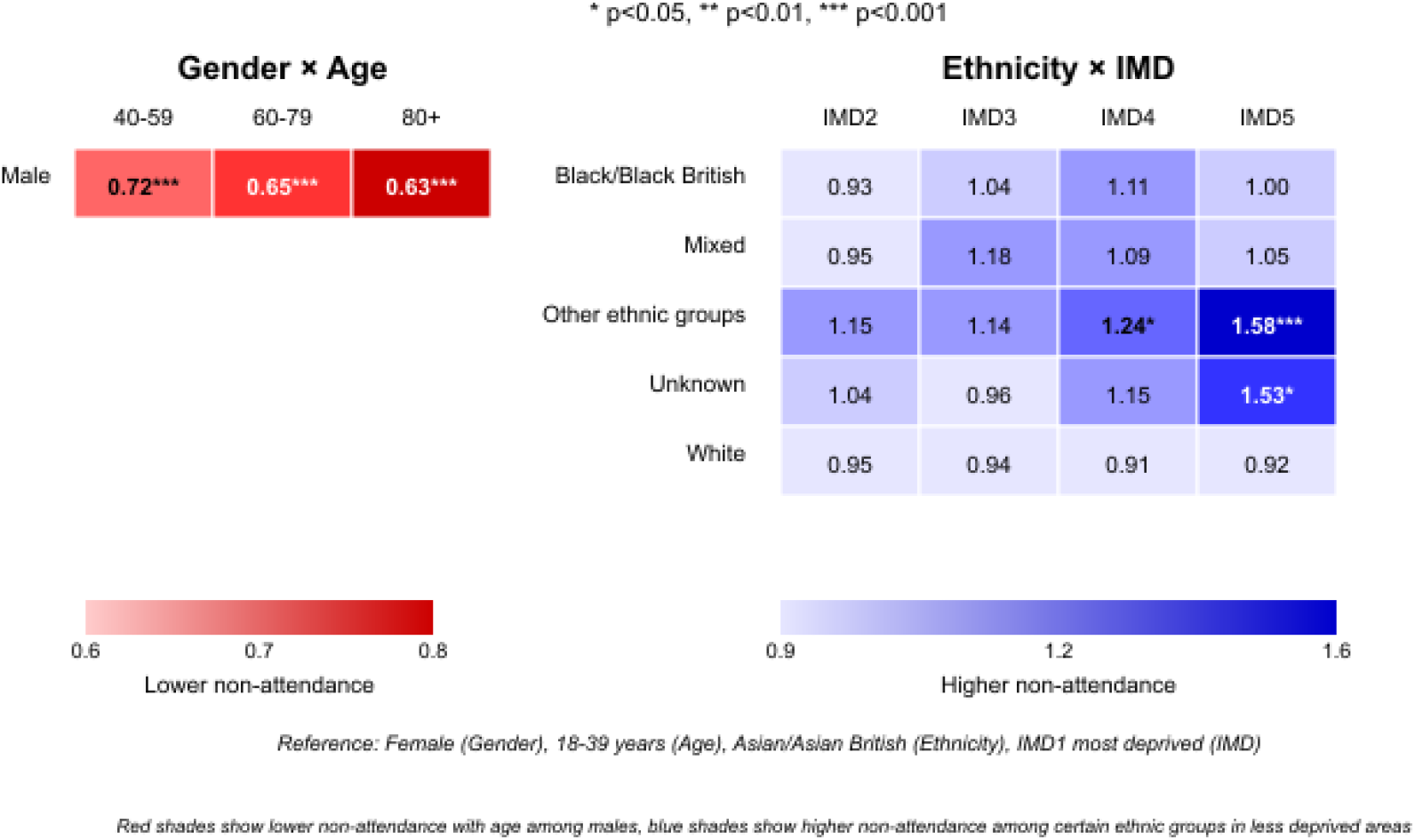
Interaction Effects on non-attendance for Gender × Age groups and Ethnicity × socioeconomic status. This heatmap presents ORs from the model interaction terms for Gender × Age (left panel) and Ethnicity × IMD (right panel). Where OR values <1 indicate lower non-attendance and values >1 indicate higher non-attendance compared to reference groups (Female, 18-39 years, Asian/Asian British, and IMD1 most deprived). The colour intensity corresponds to the magnitude of the effect: red shades (left panel) show lower non-attendance with age among males, while purple shades (right panel) show higher non-attendance among certain ethnic groups in less deprived areas, particularly “Other ethnic groups” in IMD5 areas (OR=1.58).

### Sensitivity Analyses

Propensity-score balance diagnostics demonstrated that measured covariates could be balanced between modes after weighting [NOTE: fill in SMD results from WSIC session]. Specialty-stratified analysis across 34 major specialties (those with ≥1,000 total consultations and ≥50 per mode) showed that in-person non-attendance rates exceeded telephone rates in 15 specialties (44%), while the reverse was observed in 18 (53%), with one specialty showing equal rates. The overall population-weighted pattern was driven by high-volume specialties where in-person non-attendance was substantially higher. A descriptive temporal analysis of monthly non-attendance rates by mode across the study period is presented in Supplementary Figure S1

## Discussion

### Summary of key findings

We compared non-attendance patterns between telephone and in-person secondary care outpatient consultations among adults with type 2 diabetes in Northwest London. In-person consultations had higher non-attendance than telephone consultations, across both first consultations and follow-up consultations. Key demographic factors influenced non-attendance differently by consultation mode: for in-person consultations, younger age, Black or Black British ethnicity, and greater socioeconomic deprivation were associated with higher non-attendance; for telephone consultations, male gender, younger age, “Other ethnic groups,” and greater socioeconomic deprivation were associated with higher non-attendance. The joint interaction model confirmed that demographic disparities in non-attendance were amplified for telephone consultations relative to in-person.

### Equity framing

We interpret these findings through the PROGRESS-Plus equity lens, which conceptualises health inequities along dimensions including place of residence, race/ethnicity, occupation, gender, religion, education, socioeconomic status, social capital, and the ‘Plus’ factors of age, disability, and multimorbidity. Our dataset captures several of these dimensions (place of residence via IMD quintile; ethnicity; gender; age; multimorbidity via LTC count), but does not capture education, occupation, digital literacy, or language proficiency—each of which may contribute to differential engagement with telephone care. We therefore frame observed disparities as signals of where inequity may exist, rather than exhaustive characterisations of its causes.

### Comparison with Previous Literature

Our findings contrast with some earlier research suggesting telephone care might be associated with higher non-attendance [29,30]. Instead, they align with emerging evidence that well-implemented telephone consultations can be associated with improved attendance rates [18, 19]. The demographic patterns identified in this study show important nuances in how different consultation modes affect various patient groups.

Age and gender were particularly notable. Younger age was consistently associated with higher non-attendance across both consultation modes [18, 19], with a more pronounced age gradient for telephone consultations. The gender difference in telephone attendance, with male patients showing higher non-attendance for telephone consultations, but no significant effect for in-person consultations, aligns with previous findings [19–21]. Interaction analyses further revealed that young males (18-39 years) had higher non-attendance rates compared to older age groups, suggesting this demographic may benefit from targeted intervention strategies.

Ethnic and socioeconomic differences in this study demonstrate complex patterns. The finding that Black or Black British patients had higher non-attendance for in-person consultations and ‘Other ethnic groups’ for telephone consultations suggests that cultural, linguistic, or technological barriers may manifest differently across consultation modes [22]. This aligns with recent research identifying key barriers to outpatient attendance among minority ethnic groups and those from deprived areas [23]. The socioeconomic gradient was observed for both consultation types but was notably steeper for telephone consultations, with patients from the least deprived areas (IMD5) showing lower non-attendance for telephone consultations compared to in-person. Transport barriers for patients from deprived areas likely contribute to this differential pattern of attendance (26).

The relationship between long-term conditions and attendance presented a more complex picture than previous literature suggesting uniformly better telephone attendance for patients with multiple conditions[18]. While patients with 3+ LTCs showed no significant differences in attendance patterns between consultations modes, this does not fully align with previous research that suggesting the convenience of telephone consultations particularly benefits those with complex health needs. In our study, those with 2 LTCs had significantly higher non-attendance for telephone consultations, suggesting that the convenience benefits of telephone consultations may not be evenly distributed across all patients with multiple health conditions [18, 22, 23].

Virtual care technologies may be more effective when implemented within hybrid delivery models that build upon existing in-person relationships rather than as standalone, digital-first approaches [24]. Our findings support this perspective, as we observed consistent associations with lower non-attendance across both first and follow-up consultations when delivered by telephone, suggesting that well-implemented telephone care systems may enhance patient engagement across the care continuum [25, 26].

### Strengths and limitations

This study utilised a large cohort from the WSIC database to examine attendance patterns across different demographic groups. The WSIC database includes all patients registered with general practitioners in Northweat London (around 2.7M people), and the patients in the database are representative of the local population with very limited scope for selection bias. The large sample also gives means that the study is well-powered to identify differences between population subgroups. The inclusion of both telephone and in-person consultations during the same period (January 2020 to August 2024) enabled direct comparison of attendance patterns across modalities while controlling for temporal factors that might influence healthcare utilisation. The use of zero-inflated negative binomial regression models appropriately addressed the statistical challenges of count data with overdispersion and excess zeros, providing more accurate estimates of factors associated with missed consultations.

Several limitations should be noted. First, consultations in our dataset consisted exclusively of telephone calls; our findings should not be generalised to video-based, app-based, or asynchronous digital modalities, which may show different attendance patterns and equity profiles. Second, mode assignment was non-random and the reasons for offering a telephone versus in-person consultation in any individual case are not recorded in WSIC. Although we conducted propensity-score balance diagnostics demonstrating that measured covariates can be balanced between modes, and present specialty-stratified analyses showing the pattern is broadly consistent across service lines, residual unmeasured confounding cannot be ruled out. All effect estimates should therefore be interpreted as associations rather than causal effects. Third, the primary regression model did not include specialty fixed effects or patient-level cluster-robust standard errors; the specialty-stratified analysis showed that the pattern was not uniform across all specialties, with non-attendance rates higher for telephone than in-person in 18 of 34 major specialties, indicating that specialty-level factors influence the overall finding. Formal adjustment for specialty and clustering is a priority for future work. Fourth, the study period spanned the COVID-19 pandemic and subsequent service recovery. Although a descriptive temporal analysis (Supplementary Figure S1) demonstrates that the pattern is broadly consistent across sub-periods, calendar-quarter fixed effects were not included in the primary model and time-adjusted analyses are a priority for future work. Fifth, the study was conducted in Northwest London, which has a highly diverse population and specific service configurations that may not reflect other regions or healthcare systems. Sixth, WSIC does not capture patient-level determinants of engagement such as digital literacy, language proficiency, or housing stability; these likely contribute to the ethnic and socioeconomic patterns we observed but cannot be quantified here. Finally, the ‘Other ethnic groups’ and ‘Unknown/Not stated’ categories are heterogeneous; we have retained ‘Unknown’ as a missing-indicator category, but caution is warranted when interpreting estimates for these groups.

### Implications for research, practice and policy

Future research should build upon these attendance pattern findings to explore underlying reasons for differential non-attendance, and explore barriers to attendance across modalities, particularly among patients with high non-attendance rates (older patients, Black and Mixed ethnicities). Mixed-methods studies linking mode assignment rationale to attendance outcomes would complement the population-scale findings reported here. Qualitative research can provide additional insights on patient perspectives and preferences for consultation mode. This would include looking at issues such as technological and language barriers, transportation problems or patient preferences for the type of consultation they are offered. Several research directions emerge from this study findings. First, extending these methods to video consultations and other emerging modalities will be crucial as the NHS expands its digital offerings. Second, within-patient analyses and time-adjusted models incorporating specialty fixed effects would strengthen causal inference. Third, developing dynamic algorithms that learn from local patterns could enable real-time optimisation of consultation allocation. Fourth, the high-risk profiles identified here require targeted behavioural interventions to improve engagement. Finally, validating these findings across other long-term conditions will establish broader applicability for health system transformation.

The lower non-attendance rates associated with telephone consultations, particularly for first consultations, suggest that use of telephone consultations may improve overall attendance. However, the different demographic patterns in non-attendance indicate that a personalised approach to consultation mode selection may be beneficial [27]. Patient choice could be a central consideration in this personalised approach. Offering patients the option to select their preferred consultation mode, where clinically appropriate, could enhance engagement and address inequalities, technological comfort levels, and practical constraints. Effective telephone care implementation likely includes clear communication about consultations, technical support for patients, tailored reminder systems, appropriate patient selection, and provider training. Healthcare providers should consider factors such as age, gender, ethnicity, and socioeconomic status when determining the most appropriate consultation mode [28]. They might offer technical support and additional reminders for male patients and those from “Other ethnic groups” using telephone services, while providing targeted interventions for younger patients and those from Black or Black British backgrounds attending in-person consultations. The stronger socioeconomic gradient observed for telephone consultations highlights the need for digital inclusion strategies addressing barriers to telephone care engagement, particularly for patients from deprived areas [29, 30].

Broader policy initiatives to address the digital divide are essential to ensure that the expansion of telephone and virtual care does not exacerbate existing or create new health inequalities [31]. In addition to incorporating healthcare quality performance metrics to address disparities across demographic groups and consultation modalities, policies should support digital access and literacy programmes targeted at vulnerable populations [2]. Healthcare services could implement several practical interventions to further enhance attendance, including proactive consultation reminders via SMS with detailed instructions for telephone consultations; offering brief pre-consultation technical checks for patients new to telephone care; developing digital literacy training programmes for vulnerable groups; creating a ‘buddy systems’ pairing technologically proficient patients with those less confident; and establishing dedicated telephone support lines for consultation access issues [32–34].

## Conclusion

Among adults with T2D in Northwest London, telephone consultations were associated with lower non-attendance rates across both first and follow-up secondary care specialty outpatient consultations. Attendance patterns varied across consultation modalities and were influenced by demographic factors such as age, gender, ethnicity, and socioeconomic deprivation. Younger age and greater socioeconomic deprivation consistently emerged as key predictors of non-attendance, with demographic disparities amplified for telephone relative to in-person consultations. While telephone consultations may offer opportunities to enhance engagement in secondary care, their benefits are not equally shared among all patient groups. To optimise attendance and improve continuity of care, secondary care providers should adopt a personalised, holistic approach that considers both the mode of consultation and individual patient characteristics. Whenever clinically appropriate, including patient preferences in consultation modality decisions may further enhance engagement and reduce inequalities by empowering patients to choose the consultation format that best meets their needs and circumstances. This patient-centred approach, combined with targeted interventions for groups at higher risk of non-attendance, has the potential to improve healthcare access and outcomes for people living with type 2 diabetes. These findings apply to telephone consultations and may not generalise to video-based or other digital modalities.

## Author contributions

RA: Conceptualisation, Data curation, Formal analysis, Investigation, Methodology, Software, Visualisation, Writing – original draft. ALN: Conceptualisation, Methodology, Supervision, Writing – review & editing. GK: Data curation, Methodology, Writing – review & editing. GG: Conceptualisation, Methodology, Supervision, Writing – review & editing. BH: Conceptualisation, Writing – review & editing. HK: Writing – review & editing. JV: Writing – review & editing. AM: Writing – review & editing.

## Generative AI Disclosure

Generative AI tools (including Claude by Anthropic and ChatGPT by OpenAI) were used solely for language editing and grammar polishing of text written by the authors. No content, data analysis, code, references, or scientific interpretation was generated by AI. All authors reviewed and take full responsibility for the final manuscript.

## Abbreviations

aOR: adjusted odds ratio
aRR: adjusted rate ratio
ARD: absolute risk difference
CI: confidence interval
COVID-19: coronavirus disease 2019
DNA: Did Not Attend
ED: emergency department
GP: general practitioner
IMD: Index of Multiple Deprivation
IPTW: inverse probability of treatment weighting
LTC: long-term condition
NHS: National Health Service
NIHR: National Institute for Health and Care Research
SMD: standardised mean difference
T2D: type 2 diabetes
WSIC: Whole Systems Integrated Care
ZINB: zero-inflated negative binomial

Supplementary information is available for this paper.

## Figure Legends

**Figure 1. Secondary care specialty distribution by consultation mode (In-person vs telephone).**

Bar chart showing the number of in-person (blue) and telephone (red) consultations across medical specialties for adults with type 2 diabetes in Northwest London (January 2020–August 2024). Numbers and percentages within bars represent consultation counts and proportion of total consultations for each specialty. Total consultations analysed: 850,493 (80.9% in-person, 19.1% telephone).

**Figure 2. Adjusted rate ratios (aRRs) from the count component of stratified ZINB models for nonattendance at in-person and telephone consultations.**

Adjusted rate ratios (aRRs) with 95% confidence intervals for non-attendance at in-person (blue) versus telephone (red) consultations by demographic and clinical characteristics. Reference categories are indicated in parentheses for each variable group. aRRs <1 indicate lower rate of non-attendance compared to the reference group, while aRRs >1 indicate higher rate.

**Figure 3. Interaction effects on non-attendance for Gender × Age groups and Ethnicity × socioeconomic status.**

This heatmap presents adjusted odds ratios from the within-mode interaction models for Gender × Age (left panel) and Ethnicity × IMD (right panel). Values <1 indicate lower non-attendance and values >1 indicate higher non-attendance compared to reference groups. The colour intensity corresponds to the magnitude of the effect.

