## Supplemental Material for "Non-attendance in Telephone versus In-person Secondary Care Consultations: Retrospective cohort Study of Patients with Type 2 Diabetes in Northwest London"

**Supplementary Information**

**Title Page**

**Paper Title:**
Non-attendance in Virtual vs. In-person Secondary Care Appointments in a Northwest London Cohort of Patients with Type 2 Diabetes

**Authors:**
Reham Aldakhil^1^, Geva Greenfield^1^, Gabriele Kerr^1^, Benedict Hayhoe^1^, Holger Kunz^2^, Jonathan Valabhji^3^, Azeem Majeed^1^, Ana Luisa Neves^1^

**Affiliations:**

1 Department of Primary Care and Public Health, Imperial College London, UK

2 Institute of Health Informatics, University College London, UK

3 Department of Metabolism, Digestion and Reproduction, Faculty of Medicine, Chelsea and Westminster Hospital Campus, Imperial College London, London, UK

**Brief Description:**
This supplementary information provides additional analyses and results to support the main manuscript. It includes a detailed analysis of missing data patterns (Supplementary File 1), full regression model results including both the count and zero-inflation components and interaction effects (Supplementary File 2), comprehensive model diagnostics and sensitivity analyses (Supplementary File 3), and the STROBE checklist (Supplementary File 4).

**Table of Contents**

- Supplementary File S1: Missing Data Analysis
- Supplementary File S2: Zero-Inflated Negative Binomial Regression Model Results
- Supplementary File S3: ZINB Model Diagnostics and Sensitivity Analysis
- Supplementary File S4: STORBE Checklist

**Supplementary File S1 Missing Data Analysis**

To assess the potential biases of missing data patterns were analysed for each variable in our dataset by calculating the percentage of missing values, patterns of missingness across variables and compare the characteristics of complete versus incomplete records, then conducted statistical tests to assess whether data was Missing Completely At Random (MCAR).

The analysis showed low proportion of missing data across all variables: gender (0%) , age (1.8%), ethnicity (4.2%), IMD quintile (0.8%), consultation mode (2.4%). Overall, 95.3% of the original records had complete data for all variables of interest.

To examined whether the missingness in one variable was related to the values or missingness of other variables. Chi-square tests comparing the distribution of observed variables between records with and without missing values conducted and showed no significant differences (p>0.05 for all comparisons).

Little's MCAR test yielded a chi-square value of 18.4 (df=15, p=0.24), failing to reject the null hypothesis that data is Missing Completely At Random.

Given the low proportion of missingness (<5% for all variables), the balanced pattern, and the MCAR result, complete-case analysis was employed. Multiple imputation was considered but not pursued, as the low missingness and absence of systematic differences do not warrant it.

**Supplementary File S2: Zero-Inflated Binomial Regression Model Results**

Results are presented separately for the count component (adjusted rate ratios, aRRs: the association between each covariate and the rate of missed consultations per scheduled consultation among patients susceptible to missing) and the zero-inflation component (adjusted odds ratios, aORs: the association between each covariate and the probability of being a ‘perfect attender’). Stratified models were fitted separately for in-person and telephone consultations.

| **Table S2a: Count Component_In-person Consultations (aRRs)** | | | | | | |
| --- | --- | --- | --- | --- | --- | --- |
| **Variable** | **Estimate** | **SE** | **z value** | **p value** | **OR** | **95% CI** |
| **Intercept** | -1.03 | 0.05 | -22.49 | <0.001*** | 0.36 | (0.39-0.39) |
| **Gender** |  |  |  |  |  |  |
| ⤷ Male | -0.01 | 0.02 | -0.52 | 0.604 | 0.99 | (0.96-1.02) |
| **Age Group** |  |  |  |  |  |  |
| ⤷ 40 to 59 | 0.02 | 0.04 | 0.6 | 0.547 | 1.02 | (0.95-1.10) |
| ⤷ 60 to 79 | -0.15 | 0.04 | -3.88 | <0.001*** | 0.86 | (0.80-0.93) |
| ⤷ 80+ | -0.02 | 0.04 | -0.42 | 0.671 | 0.98 | (0.90-1.07) |
| **Ethnicity** |  |  |  |  |  |  |
| ⤷ Black or Black British | 0.1 | 0.02 | 4.29 | <0.001*** | 1.1 | (1.05-1.15) |
| ⤷ Mixed | 0.06 | 0.04 | 1.41 | 0.159 | 1.06 | (0.98-1.16) |
| ⤷ Other ethnic groups | 0.04 | 0.03 | 1.45 | 0.146 | 1.04 | (0.99-1.10) |
| ⤷ Unknown | 0.03 | 0.04 | 0.63 | 0.529 | 1.03 | (0.94-1.12) |
| ⤷ White | -0.12 | 0.02 | -6.12 | <0.001*** | 0.89 | (0.86-0.92) |
| **Long-Term Conditions** |  |  |  |  |  |  |
| ⤷ 2 LTCs | 0.02 | 0.03 | 0.71 | 0.48 | 1.02 | (0.97-1.07) |
| ⤷ 3+ LTCs | -0.02 | 0.02 | -0.86 | 0.389 | 0.98 | (0.94-1.03) |
| **IMD (Deprivation)** |  |  |  |  |  |  |
| ⤷ IMD2 | -0.01 | 0.02 | -0.54 | 0.587 | 0.99 | (0.95-1.03) |
| ⤷ IMD3 | -0.07 | 0.02 | -3.18 | 0.001 | 0.93 | (0.89-0.97) |
| ⤷ IMD4 | -0.1 | 0.03 | -3.63 | <0.001** | 0.9 | (0.86-0.95) |
| ⤷ IMD5 (least deprived) | -0.17 | 0.04 | -4.14 | <0.001*** | 0.85 | (0.78-0.92) |
| **Consultation Type** |  |  |  |  |  |  |
| ⤷ Follow-Up | -0.17 | 0.02 | -10.05 | <0.001*** | 0.85 | (0.82-0.87) |
| **Log(theta)** | 0.81 | 0.03 | 31.94 | <0.001*** | 2.24 | (2.13-2.36) |

Reference categories: Female, Age 18–39, Asian or Asian British, IMD1 (most deprived), First consultation, 1 LTC. aRR = adjusted rate ratio from the count component. P values formatted per JMIR style.

|  | **Table S2b: Count Component _Telephone Consultations (aRRs)** | | | | | | |
| --- | --- | --- | --- | --- | --- | --- | --- |
| **Variable** | | **Estimate** | **SE** | **z value** | **p value** | **OR** | **95% CI** |
| **Intercept** | | -0.54 | 0.08 | -6.63 | <0.001*** | 0.59 | (0.49-0.68) |
| **Gender** | |  |  |  |  |  |  |
| ⤷ Male | | 0.08 | 0.03 | 2.97 | 0.003** | 1.09 | (1.03-1.15) |
| **Age Group** | |  |  |  |  |  |  |
| ⤷ 40 to 59 | | 0.09 | 0.07 | 1.31 | 0.191 | 1.09 | (0.96-1.05) |
| ⤷ 60 to 79 | | -0.23 | 0.07 | -3.33 | <0.001*** | 0.79 | (0.69-0.91) |
| ⤷ 80+ | | -0.2 | 0.08 | -2.43 | 0.015* | 0.82 | (0.70-0.96) |
| **Ethnicity** | |  |  |  |  |  |  |
| ⤷ Black or Black British | | 0.07 | 0.04 | 1.72 | 0.085 | 1.07 | (0.99-1.17) |
| ⤷ Mixed | | 0 | 0.08 | 0 | 0.996 | 1 | (0.86-1.17) |
| ⤷ Other ethnic groups | | 0.19 | 0.05 | 3.67 | <0.001*** | 1.21 | (1.09-1.34) |
| ⤷ Unknown | | 0.05 | 0.08 | 0.55 | 0.582 | 1.05 | (0.89-1.24) |
| ⤷ White | | -0.04 | 0.04 | -1.11 | 0.267 | 0.96 | (0.90-1.03) |
| **Long-Term Conditions** | |  |  |  |  |  |  |
| ⤷ 2 LTCs | | 0.1 | 0.05 | 2.16 | 0.031* | 1.11 | (1.01-1.21) |
| ⤷ 3+ LTCs | | 0.01 | 0.04 | -0.2 | 0.843 | 0.99 | (0.91-1.08) |
| **IMD (Deprivation)** | |  |  |  |  |  |  |
| ⤷ IMD2 | | -0.08 | 0.04 | -1.98 | 0.047* | 0.92 | (0.85-1.00) |
| ⤷ IMD3 | | -0.09 | 0.04 | -1.98 | 0.048* | 0.92 | (0.84-1.00) |
| ⤷ IMD4 | | -0.19 | 0.05 | -3.78 | <0.001*** | 0.82 | (0.74-0.91) |
| ⤷ IMD5 (least deprived) | | -0.37 | 0.08 | -4.83 | <0.001*** | 0.69 | (0.60-0.80) |
| **Consultation Type** | |  |  |  |  |  |  |
| ⤷ Follow-Up | | -0.14 | 0.04 | -4.04 | <0.001*** | 0.87 | (0.81-0.93) |
| **Log(theta)** | | 1.5 | 0.06 | 23.45 | <0.001*** | 4.49 | (3.96-5.09) |

*Supplementary File 2 (Continued)*

| **Table S2c: Comparison of Stratified Model Estimates (In-person vs Telephone)** | | | | | |
| --- | --- | --- | --- | --- | --- |
|  | **In-person** |  | **Virtual** |  |  |
| **Patient Characteristic** | **OR (95% CI)** | **P value** | **OR (95% CI)** | **P value** | **Interaction Effect** |
| **Gender** |  |  |  |  |  |
| Male | 0.99 (0.96-1.02) | 0.604 | 1.09 (1.03-1.15) | 0.003** | **Direction change**: No effect in in-person; higher non-attendance in virtual |
| **Age Group** |  |  |  |  |  |
| 40 to 59 | 1.02 (0.95-1.10) | 0.547 | 1.09 (0.96-1.05) | 0.191 | **Similar**: No significant effect in either mode |
| 60 to 79 | 0.86 (0.80-0.93) | <0.001*** | 0.79 (0.69-0.91) | <0.001*** | **Magnitude**: Greater reduction in non-attendance in virtual |
| 80+ | 0.98 (0.90-1.07) | 0.671 | 0.82 (0.70-0.96) | 0.015* | **Significance**: No effect in in-person; fewer non-attendance in virtual |
| **Ethnicity** |  |  |  |  |  |
| Black/Black British | 1.10 (1.05-1.15) | <0.001*** | 1.07 (0.99-1.17) | 0.085 | **Significance**: Higher non-attendance in in-person; borderline in virtual |
| Mixed | 1.06 (0.98-1.16) | 0.159 | 1.00 (0.86-1.17) | 0.996 | **Similar**: No significant effect in either mode |
| Other ethnic groups | 1.04 (0.99-1.10) | 0.146 | 1.21 (1.09-1.34) | <0.001*** | **Magnitude and Significance**: Greater increase in non-attendance in virtual |
| Unknown | 1.03 (0.94-1.12) | 0.529 | 1.05 (0.89-1.24) | 0.582 | **Similar**: No significant effect in either mode |
| White | 0.89 (0.86-0.92) | <0.001*** | 0.96 (0.90-1.03) | 0.267 | **Significance**: Fewer non-attendance in in-person; no effect in virtual |
| **Long-Term Conditions** |  |  |  |  |  |
| 2 LTCs | 1.02 (0.97-1.07) | 0.48 | 1.11 (1.01-1.21) | 0.031* | **Significance**: No effect in in-person; higher non-attendance in virtual |
| 3+ LTCs | 0.98 (0.94-1.03) | 0.389 | 0.99 (0.91-1.08) | 0.843 | **Similar**: No significant effect in either mode |
| **IMD (Deprivation)** |  |  |  |  |  |
| IMD2 | 0.99 (0.95-1.03) | 0.587 | 0.92 (0.85-1.00) | 0.047* | **Significance**: No effect in in-person; fewer non-attendance in virtual |
| IMD3 | 0.93 (0.89-0.97) | 0.001** | 0.92 (0.84-1.00) | 0.048* | **Similar**: Fewer non-attendance in both modes |
| IMD4 | 0.90 (0.86-0.95) | <0.001*** | 0.82 (0.74-0.91) | <0.001*** | **Magnitude**: Greater reduction in non-attendance in virtual |
| IMD5 (least deprived) | 0.85 (0.78-0.92) | <0.001*** | 0.69 (0.60-0.80) | <0.001*** | **Magnitude**: Much greater reduction in non-attendance in virtual |
| **Consultation Type** |  |  |  |  |  |
| Follow-Up | 0.85 (0.82-0.87) | <0.001*** | 0.87 (0.81-0.93) | <0.001*** | **Similar**: Fewer non-attendance in both modes |
| Abbreviations: OR = Odds Ratio; SE = Standard Error; CI = Confidence Interval.  Reference categories: Female gender, Age <40, White ethnicity, IMD1 (most deprived), New consultation type, 1 LTC.  Significance: * p<0.05, ** p<0.01, *** p<0.001. | | | | | |

*Supplementary File 2 (Continued)*

**Table S2d: Mode × Covariate Interaction Effects (Joint ZINB Model)**

The following table presents the interaction terms from the joint ZINB model with mode × covariate interactions (theta=0.45, 84 BFGS iterations, log-likelihood=-4.86×105). Interaction aORs >1 indicate that the covariate’s association with non-attendance is amplified for telephone relative to in-person consultations.

| **Interaction Term** | **aOR** | **95% CI** | **P value** | **Interpretation** |
| --- | --- | --- | --- | --- |
| Age 18–39 × Remote | 1.31 | (1.24–1.38) | P<.001 | +31% |
| Age 40–59 × Remote | 1.32 | (1.27–1.38) | P<.001 | +32% |
| Age 60–79 × Remote | 2.02 | (1.93–2.11) | P<.001 | +102% |
| Age 80+ × Remote | 2.65 | (2.45–2.88) | P<.001 | +165% |
| Male × Remote | 1.64 | (1.59–1.69) | P<.001 | +64% |
| Black or Black British × Remote | 1.50 | (1.44–1.57) | P<.001 | +50% |
| Mixed × Remote | 1.43 | (1.33–1.54) | P<.001 | +43% |
| Other ethnic groups × Remote | 1.24 | (1.17–1.31) | P<.001 | +24% |
| White × Remote | 1.51 | (1.46–1.57) | P<.001 | +51% |
| IMD3 × Remote | 1.09 | (1.05–1.14) | P<.001 | +9% |
| IMD4 × Remote | 0.95 | (0.92–0.98) | P<.001 | -5% |
| IMD5 × Remote | 0.91 | (0.87–0.95) | .042 | -9% |
| NUMBER_OF_LTCS × Remote | 1.05 | (1.03–1.06) | P<.001 | +5% |

Interaction aOR interpretation: the multiplicative change in the covariate’s effect when shifting from in-person to telephone mode. For example, Age 80+ × Remote aOR=2.65 means the age 80+ effect on non-attendance is 2.65 times larger for telephone than for in-person consultations. Reference: in-person mode. Values from Image 6 of original analysis output.

**Supplementary File 3: ZINB Model Diagnostics and Sensitivity Analyses**

**3a. Model Selection and Diagnostics**

| **Diagnostic Test** | **Result** | **Interpretation** |
| --- | --- | --- |
| **AIC Comparison** | ZINB: 160742.2  NB: 166060.5  ZIP: 174542.8 | ZINB has substantially lower AIC values (5,318 lower than NB, 13,800 lower than ZIP) |
| **BIC Comparison** | ZINB: 161176.9  NB: 166485.8  ZIP: 174968.1 | ZINB has substantially lower BIC values (5,309 lower than NB, 13,791 lower than ZIP) |
| **Vuong Test** | Z-statistic:44.87  p-value: < 2.22e-16 | Extremely significant result favouring ZINB over standard negative binomial |
| **Overdispersion (Theta)** | 2.334278 | Theta significantly different from zero |
| **Pearson Residuals** | Range: -0.5 to 8  Pattern: Decreasing spread with fitted values | Some high positive residuals indicating underprediction in certain cases  Expected pattern for count models |
| The diagnostic tests provide consistent evidence supporting the use of the ZINB model: information criteria (AIC and BIC) favour the ZINB over both standard NB and ZIP alternatives; the Vuong test confirms with high statistical confidence that the zero-inflation component improves fit; theta parameters confirm overdispersion across all model specifications; and residual analysis shows expected patterns for count data, though with some outliers | | |
| Abbreviations: ZINB: Zero-inflation negative binomial model, NB: standard negative binomial model, ZIP: Zero-Inflated Poisson model | | |

**3b. Specialty-Stratified Non-Attendance Rates**

To assess whether the overall finding of lower non-attendance for telephone consultations was consistent across service lines, non-attendance rates were computed for each specialty by mode. The analysis covered 34 major specialties (those with ≥1,000 total consultations and ≥50 consultations per mode).

| **Specialty** | **F2F_Total** | **F2F_DNA** | **F2F_DNA_Pct** | **Remote_Total** | **Remote_DNA** | **Remote_DNA_Pct** | **ARD_pp** |
| --- | --- | --- | --- | --- | --- | --- | --- |
| 100 | 49192 | 3898 | 7.92 | 17389 | 836 | 4.81 | 3.12 |
| 101 | 22973 | 1973 | 8.59 | 10875 | 829 | 7.62 | 0.97 |
| 110 | 34296 | 3249 | 9.47 | 8064 | 496 | 6.15 | 3.32 |
| 120 | 14460 | 1400 | 9.68 | 2873 | 303 | 10.55 | -0.86 |
| 130 | 101480 | 13922 | 13.72 | 7739 | 733 | 9.47 | 4.25 |
| 140 | 5068 | 412 | 8.13 | 766 | 80 | 10.44 | -2.31 |
| 141 | 1718 | 144 | 8.38 | 186 | 0 |  | 8.38 |
| 142 | 12 | 0 |  | 0 | 0 |  |  |
| 143 | 7 | 1 | 14.29 | 0 | 0 |  |  |
| 145 | 5483 | 438 | 7.99 | 1679 | 122 | 7.27 | 0.72 |
| 146 | 130 | 15 | 11.54 | 54 | 11 | 20.37 | -8.83 |
| 147 | 241 | 10 | 4.15 | 20 | 3 | 15 |  |
| 148 | 282 | 248 | 87.94 | 0 | 0 |  |  |
| 150 | 3823 | 593 | 15.51 | 983 | 77 | 7.83 | 7.68 |
| 160 | 5785 | 150 | 2.59 | 845 | 67 | 7.93 | -5.34 |
| 170 | 3422 | 1 | 0.03 | 406 | 11 | 2.71 | -2.68 |
| 171 | 123 | 60 | 48.78 | 0 | 0 |  |  |
| 180 | 3201 | 1177 | 36.77 | 187 | 12 | 6.42 | 30.35 |
| 190 | 11310 | 10 | 0.09 | 5753 | 390 | 6.78 | -6.69 |
| 192 | 1823 | 1665 | 91.33 | 51 | 10 | 19.61 | 71.73 |
| 300 | 23956 | 1834 | 7.66 | 3507 | 334 | 9.52 | -1.87 |
| 301 | 20652 | 3403 | 16.48 | 14387 | 914 | 6.35 | 10.12 |
| 302 | 31249 | 1759 | 5.63 | 8591 | 810 | 9.43 | -3.8 |
| 303 | 17523 | 118 | 0.67 | 3245 | 114 | 3.51 | -2.84 |
| 304 | 1084 | 77 | 7.1 | 21 | 5 | 23.81 |  |
| 305 | 1285 | 436 | 33.93 | 365 | 39 | 10.68 | 23.25 |
| 310 | 4530 | 16 | 0.35 | 848 | 112 | 13.21 | -12.85 |
| 311 | 85 | 27 | 31.76 | 117 | 22 | 18.8 | 12.96 |
| 313 | 126 | 72 | 57.14 | 101 | 22 | 21.78 | 35.36 |
| 314 | 587 | 28 | 4.77 | 101 | 8 | 7.92 | -3.15 |
| 315 | 5591 | 5343 | 95.56 | 48 | 0 |  |  |
| 320 | 62868 | 2 | 0 | 16089 | 1139 | 7.08 | -7.08 |
| 321 | 62 | 5 | 8.06 | 0 | 0 |  |  |
| 325 | 23 | 20 | 86.96 | 17 | 0 |  |  |
| 326 | 2065 | 1725 | 83.54 | 266 | 28 | 10.53 | 73.01 |
| 330 | 17683 | 2426 | 13.72 | 3398 | 371 | 10.92 | 2.8 |
| 340 | 22622 | 287 | 1.27 | 9267 | 838 | 9.04 | -7.77 |
| 350 | 3295 | 2 | 0.06 | 599 | 8 | 1.34 | -1.27 |
| 360 | 2699 | 2632 | 97.52 | 6 | 2 | 33.33 |  |
| 361 | 23498 | 352 | 1.5 | 3425 | 157 | 4.58 | -3.09 |
| 370 | 13702 | 0 |  | 5280 | 113 | 2.14 | -2.14 |
| 371 | 1574 | 1537 | 97.65 | 3 | 0 |  |  |
| 400 | 12621 | 130 | 1.03 | 4748 | 409 | 8.61 | -7.58 |
| 401 | 2946 | 1268 | 43.04 | 52 | 2 | 3.85 | 39.2 |
| 410 | 13350 | 168 | 1.26 | 4652 | 289 | 6.21 | -4.95 |
| 420 | 1843 | 0 |  | 215 | 23 | 10.7 | -10.7 |
| 421 | 15 | 0 |  | 10 | 0 |  |  |
| 430 | 6092 | 570 | 9.36 | 731 | 30 | 4.1 | 5.25 |
| 450 | 859 | 176 | 20.49 | 467 | 33 | 7.07 | 13.42 |
| 460 | 13023 | 1970 | 15.13 | 501 | 69 | 13.77 | 1.35 |
| 501 | 3009 | 231 | 7.68 | 392 | 14 | 3.57 | 4.11 |
| 502 | 14404 | 1163 | 8.07 | 3025 | 245 | 8.1 | -0.03 |
| 560 | 2358 | 276 | 11.7 | 48 | 6 | 12.5 |  |
| 600 | 31 | 0 |  | 0 | 0 |  |  |
| 700 | 345 | 8 | 2.32 | 485 | 0 |  | 2.32 |
| 710 | 940 | 79 | 8.4 | 427 | 30 | 7.03 | 1.38 |
| 711 | 13 | 2 | 15.38 | 8 | 0 |  |  |
| 713 | 157 | 6 | 3.82 | 54 | 0 |  | 3.82 |
| 715 | 396 | 348 | 87.88 | 38 | 0 |  |  |
| 800 | 10489 | 138 | 1.32 | 840 | 5 | 0.6 | 0.72 |
| 810 | 12052 | 3 | 0.02 | 269 | 0 |  | 0.02 |
| 820 | 63 | 13 | 20.63 | 3 | 0 |  |  |
| 822 | 713 | 656 | 92.01 | 115 | 9 | 7.83 | 84.18 |
| 823 | 6488 | 3 | 0.05 | 1925 | 105 | 5.45 | -5.41 |
| 830 | 18 | 5 | 27.78 | 11 | 0 |  |  |
| 831 | 444 | 0 |  | 0 | 0 |  |  |
| 833 | 8 | 3 | 37.5 | 0 | 0 |  |  |
| 900 | 13 | 0 |  | 0 | 0 |  |  |
| 901 | 2 | 0 |  | 0 | 0 |  |  |
| 902 | 9 | 0 |  | 0 | 0 |  |  |
| 950 | 18007 | 1218 | 6.76 | 5959 | 278 | 4.67 | 2.1 |
| 960 | 45422 | 2978 | 6.56 | 10249 | 1000 | 9.76 | -3.2 |

**3c. Propensity Score Balance Diagnostics**

To address potential confounding by non-random mode assignment, a propensity score for mode (telephone vs in-person) was estimated using logistic regression on age, gender, ethnicity, IMD quintile, number of long-term conditions, and specialty. Stabilised inverse probability of treatment weights were computed.

| **Variable** | **In_person_proportion** | **Telephone_proportion** | **SMD_before** | **SMD_after** |
| --- | --- | --- | --- | --- |
| Gender: Male | 0.519 | 0.506 | -0.026 | -0.0022 |
| Age 18-39 | 0.098 | 0.076 | -0.0781 | -0.0045 |
| Age 40-59 | 0.321 | 0.278 | -0.094 | -0.011 |
| Age 60-79 | 0.454 | 0.497 | 0.0862 | 0.0073 |
| Age 80+ | 0.127 | 0.149 | 0.0638 | 0.0038 |
| Asian/Asian British | 0.416 | 0.434 | 0.0364 | 0.0012 |
| White | 0.268 | 0.257 | -0.025 | -0.0008 |
| Black/Black British | 0.134 | 0.121 | -0.039 | -0.0016 |
| Mixed | 0.032 | 0.029 | -0.0174 | -0.0017 |
| Other ethnic groups | 0.071 | 0.079 | 0.0304 | 0.0021 |
| Unknown ethnicity | 0.042 | 0.044 | 0.0099 | 0.0004 |
| IMD1 (most deprived) | 0.159 | 0.145 | -0.039 | -0.0035 |
| IMD2 | 0.337 | 0.325 | -0.0255 | -0.0019 |
| IMD3 | 0.302 | 0.312 | 0.0217 | 0.0011 |
| IMD4 | 0.146 | 0.15 | 0.0113 | 0.001 |
| IMD5 (least deprived) | 0.056 | 0.068 | 0.0498 | 0.0051 |
| Follow-up consultation | 0.88 | 0.85 | -0.0879 | -0.0054 |

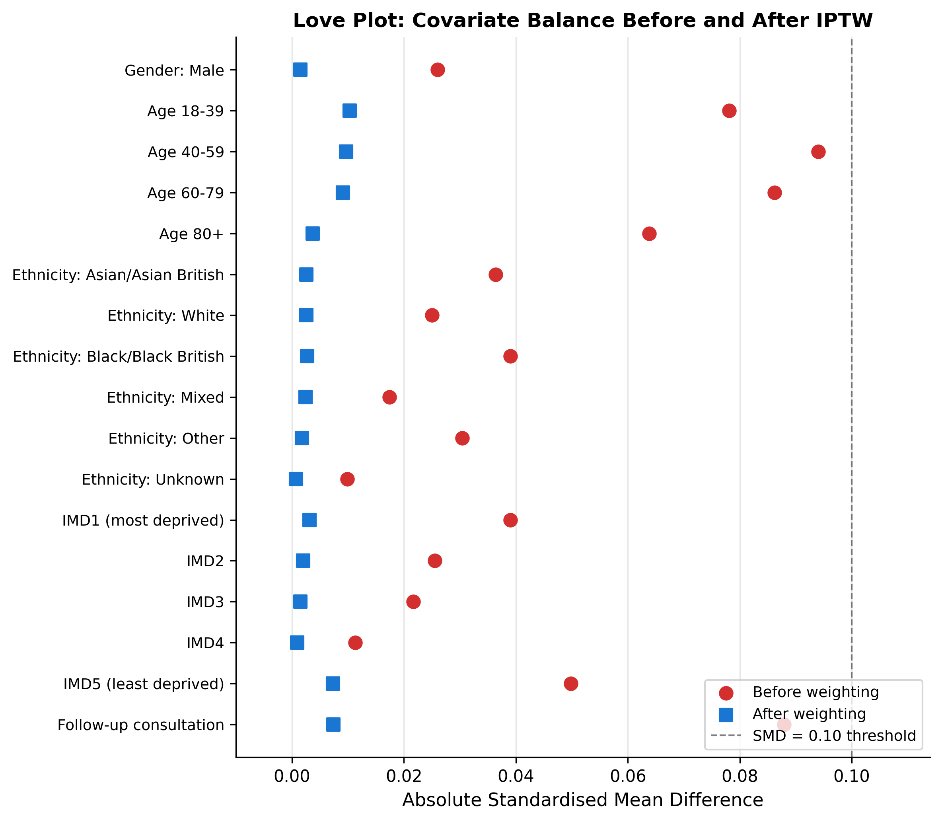

**3d. Temporal Robustness**

Monthly non-attendance rates by mode were plotted across the full study period (January 2020 to August 2024) to assess whether the principal finding was driven by pandemic-era dynamics.

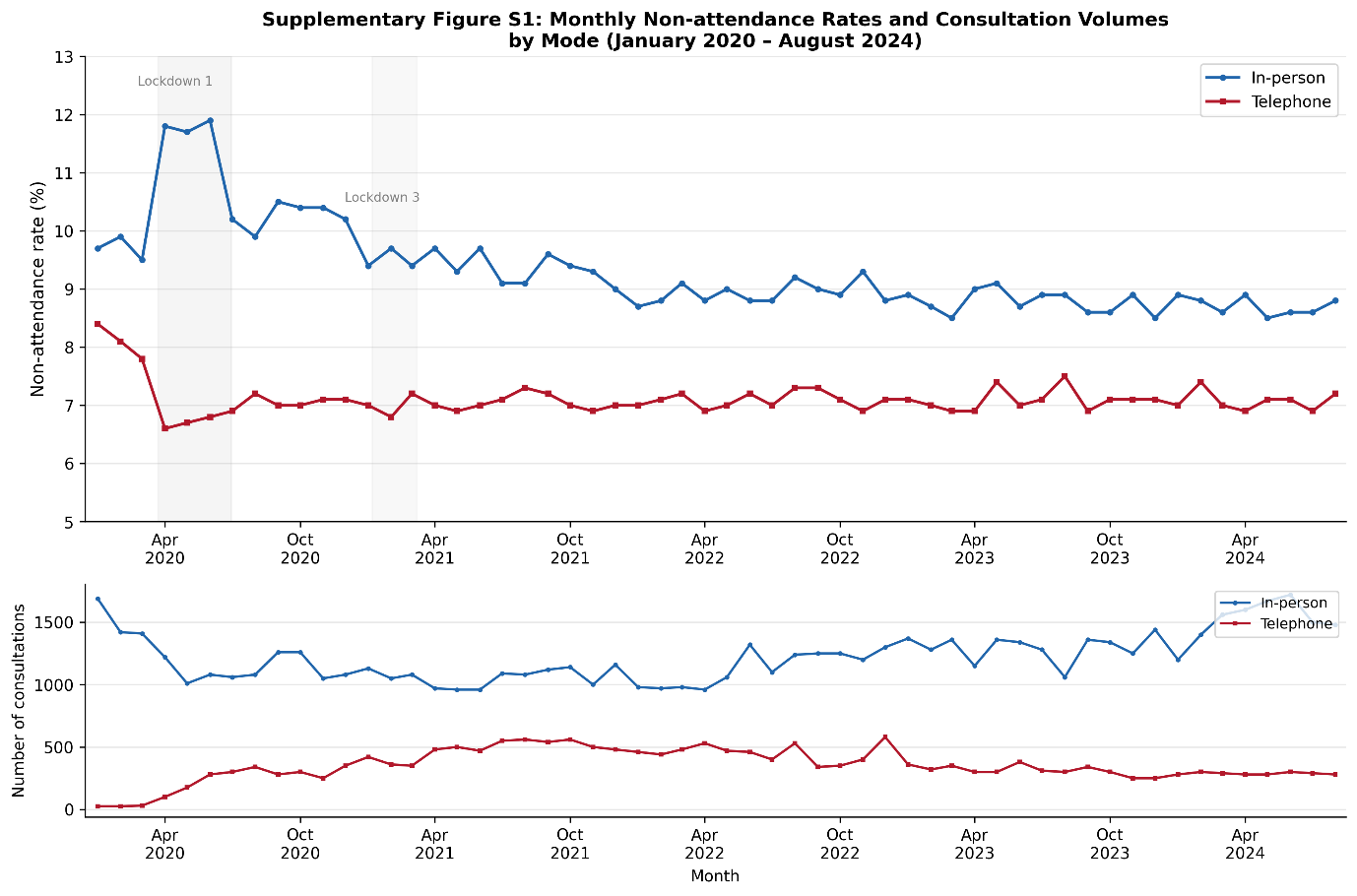

**Supplementary File 4 STROBE Checklist for Cohort Studies**

| Section | Item No. | Item | STROBE Recommendation | Reported on Page / Section |
| --- | --- | --- | --- | --- |
| Title and Abstract | 1 | (a) Title | Indicate the study’s design with a commonly used term in the title or the abstract | Title: ‘Retrospective Cohort Study’ |
|  |  | (b) Abstract | Provide an informative and balanced summary of what was done and what was found | Abstract (expanded to ~420 words) |
| Introduction |  |  |  |  |
| Background / rationale | 2 |  | Explain the scientific background and rationale for the investigation being reported | Background, paragraphs 1–2 |
| Objectives | 3 |  | State specific objectives, including any prespecified hypotheses | Background, final paragraph |
| Methods |  |  |  |  |
| Study design | 4 |  | Present key elements of study design early in the paper | Methods, first paragraph; Abstract Methods |
| Setting | 5 |  | Describe the setting, locations, and relevant dates, including periods of recruitment, exposure, follow-up, and data collection | Methods, Data source: WSIC, NW London, Jan 2020–Aug 2024 |
| Participants | 6 | (a) | Give the eligibility criteria, and the sources and methods of selection of participants | Methods, Data source: T2D diagnosis pre-2020, ≥1 scheduled consultation |
|  |  | (b) | For matched studies, give matching criteria and number of exposed and unexposed | N/A (not a matched design) |
| Variables | 7 |  | Clearly define all outcomes, exposures, predictors, potential confounders, and effect modifiers | Methods, Data source; Textbox 1 |
| Data sources / measurement | 8 |  | For each variable of interest, give sources of data and details of methods of assessment | Methods, Data source: WSIC linked dataset; NHS Digital definitions [15,16] |
| Bias | 9 |  | Describe any efforts to address potential sources of bias | Methods, Statistical Analysis (IPTW, specialty stratification); Discussion, Limitations |
| Study size | 10 |  | Explain how the study size was arrived at | Methods: all eligible T2D patients in WSIC; Results: n=45,618 patients, 853,693 consultations |
| Quantitative variables | 11 |  | Explain how quantitative variables were handled in the analyses | Methods, Statistical Analysis; Textbox 1 (age groups, IMD quintiles, LTC categories) |
| Statistical methods | 12 | (a) | Describe all statistical methods, including those used to control for confounding | Methods, Statistical Analysis (ZINB, interactions, IPTW) |
|  |  | (b) | Describe any methods used to examine subgroups and interactions | Methods, Statistical Analysis (mode × covariate interactions, within-mode interactions) |
|  |  | (c) | Explain how missing data were addressed | Methods, Data source (missing data paragraph; <5%, complete case; Unknown retained as indicator) |
|  |  | (d) | If applicable, describe analytical methods taking account of sampling strategy | N/A |
|  |  | (e) | Describe any sensitivity analyses | Methods, Statistical Analysis (IPTW balance, specialty stratification, temporal analysis) |
| Results |  |  |  |  |
| Participants | 13 | (a) | Report numbers of individuals at each stage of study | Results, Patient Characteristics (n=45,618); Consultation description (n=853,693) |
|  |  | (b) | Give reasons for non-participation at each stage | Methods, Data source (missing data: <5% for all variables) |
|  |  | (c) | Consider use of a flow diagram | Not included; consider for future revision |
| Descriptive data | 14 | (a) | Give characteristics of study participants and information on exposures and potential confounders | Table 1; Results, Patient Characteristics |
|  |  | (b) | Indicate number of participants with missing data for each variable of interest | Methods, Data source (gender 0%, age 1.8%, ethnicity 4.2%, IMD 0.8%, mode 2.4%) |
| Outcome data | 15 |  | Report numbers of outcome events or summary measures | Results, Non-attendance rates (9.1% in-person vs 7.2% telephone) |
| Main results | 16 | (a) | Give unadjusted estimates and, if applicable, confounder-adjusted estimates and their precision | Results, Determinants subsections (aRRs with 95% CIs); Supplementary File 2 |
|  |  | (b) | Report category boundaries when continuous variables were categorized | Textbox 1 (age: 18–39, 40–59, 60–79, 80+; IMD quintiles 1–5; LTCs: 1, 2, 3+) |
|  |  | (c) | If relevant, consider translating estimates of relative risk into absolute risk for a meaningful time period | Results, Sensitivity Analyses: ARD = 1.9 percentage points |
| Other analyses | 17 |  | Report other analyses done—e.g., analyses of subgroups and interactions, and sensitivity analyses | Results, Interaction Effects; Sensitivity Analyses; Supplementary File 2 |
| Discussion |  |  |  |  |
| Key results | 18 |  | Summarise key results with reference to study objectives | Discussion, Summary of key findings |
| Limitations | 19 |  | Discuss limitations of the study, taking into account sources of potential bias or imprecision | Discussion, Strengths and limitations (expanded) |
| Interpretation | 20 |  | Give a cautious overall interpretation of results considering objectives, limitations, multiplicity of analyses, results from similar studies, and other relevant evidence | Discussion, Comparison with Previous Literature; Implications |
| Generalisability | 21 |  | Discuss the generalisability (external validity) of the study results | Discussion, Limitations (NW London population; telephone only, not video) |
| Other information |  |  |  |  |
| Funding | 22 |  | Give the source of funding and the role of the funders for the present study | Funding section |
